# Use of the International Classification of Diseases to Perinatal Mortality (ICD-PM) with verbal autopsy to determine the causes of stillbirths and neonatal deaths in rural Cambodia: a population-based, prospective, cohort study

**DOI:** 10.1101/2025.05.21.25328109

**Authors:** Kaajal Patel, Sopheakneary Say, Daly Leng, Sophanou Khut, Sothearith Duong, Chou Ly, Arthur Riedel, Koung Lo, Verena Carrara, Claudia Turner

## Abstract

**Background:** Perinatal mortality remains a significant global health challenge, particularly in low- and middle-income countries (LMICs). Accurate cause-of-death data are essential to inform effective interventions but is often scarce. This study aimed to identify causes of stillbirths and neonatal deaths in rural Cambodia using verbal autopsy (VA) and the WHO International Classification of Diseases to Perinatal Mortality (ICD-PM).

**Methods:** A four-year prospective study (2018-2022) in Preah Vihear province, Cambodia, established a community health worker-based pregnancy surveillance system. Verbal autopsy was conducted on stillbirths and neonatal deaths, with dual physician analysis to interpret VA data. To classify causes of death, ICD-PM was applied with adaptations made for stillbirths with unknown timing of death.

**Results:** A total of 522 deaths (229 stillbirths, 293 neonatal deaths) were recorded, and 79.1% (413) had a VA. Applying ICD-PM, primary causes of death were identified for 36.6% of stillbirths and 95.0% of neonatal deaths. The leading cause of death was hypoxia for intrapartum stillbirths (78.3%), low birth weight and prematurity for early neonatal deaths (40.9%), and infection for late neonatal deaths (51.4%). Complications during labour and delivery were the leading maternal contributing condition for intrapartum stillbirths (63.3%) and early neonatal deaths (42.4%). Unknown timing of death was assigned to 12.0% of stillbirths.

**Conclusion:** Application of ICD-PM with VA-derived data provides valuable insights into causes of stillbirths and neonatal deaths. However, adaptations are necessary to address ICD-PM’s limitations, particularly to classify unknown timing of death. Our findings can contribute to global efforts to improve the reporting of perinatal mortality data.

## Introduction

Despite increased global focus and investment, perinatal mortality remains unacceptably high, amounting to 2.3 million neonatal deaths a year globally, with low- and middle-income countries (LMICs) carrying the vast burden.^1^ To address this urgent global health challenge, population-level perinatal mortality data are needed to inform programmes and policy, particularly given limited resources. Knowledge of how many, who, where, and why deaths are occurring allows for targeted interventions, especially important for perinatal deaths, which are mostly preventable with available, inexpensive interventions.^2^

Perinatal mortality data comprise mortality rates, as well as timing and causes of deaths. To be meaningful, cause of death data need to be accurate, reliable, feasible, relevant, and comparable across different settings. However, in areas with the highest perinatal mortality, the quality of data is poorest: less than 3% of perinatal deaths in LMICs achieve complete recording and certification^3^ compared to more than 90% in high-income countries.^4^

A civil registration and vital statistics (CRVS) system is the international gold standard for generating reliable, comprehensive, and timely statistics on all births and deaths in a population. Well-functioning CRVS systems are associated with better health outcomes.^5^ Despite it’s importance, emphasised by a Sustainable Development Goal focusing on improving CRVS,^6^ CRVS systems remain weak in many LMICs, resulting in poor quality perinatal mortality rate and cause of death data. This is for two main reasons. Firstly, many births and especially deaths occur without contact with the health system, resulting in millions of births and deaths never being officially recorded, and referred to as a ‘scandal of invisibility’.^7,8^ Secondly, weak health systems can mean facility-based births and deaths remain unrecorded, records are lost, and/or causes of deaths are inaccurate.

While CRVS systems in Cambodia are still developing, perinatal mortality rate estimates are available from demographic health surveys (DHS).^9,10^ However, the Cambodian DHS lacks data on timing of stillbirths (antepartum or intrapartum) and causes of perinatal deaths. Data on perinatal causes of death in Cambodia are scarce, derived mainly from small studies and estimates.^11–14^ The limited availability and quality of these data necessitate reliance on estimates in perinatal epidemiology.^15,16^ However, there is increasing recognition that direct measurement, not estimation, is essential for accurate mortality data.^17^

In low-resource settings, robust yet feasible interim solutions are needed to provide mortality data until CRVS systems are strengthened. Surveillance and verbal autopsy (VA) are prominent strategies to find births and deaths and determine causes of deaths respectively.^18^ Population-based pregnancy surveillance systems aim to prospectively identify all relevant cases and follow them up for a defined time period to establish pregnancy outcomes and collect relevant data. Verbal autopsy involves data collection on the events preceding death, collected via interviews with families of the deceased, and then data analysis to determine the cause of death for each case.

Perhaps surprisingly given the necessity of consistent and comparable data to tackle perinatal mortality, a standardised global classification system to define perinatal causes of deaths was only introduced in 2016: the WHO application of ICD-10 to perinatal deaths: the ICD-perinatal mortality classification system (ICD-PM).^19^ ICD-PM uses a three-step approach to first determine the timing of death (antepartum, intrapartum, or postpartum/neonatal), second determine the primary medical cause of death, and third determine the contributing maternal condition.^19,20^

A recent systematic review found that ICD-PM had been applied in low-income settings to only 6% (2556/45,735) of perinatal death cases, which exclusively comprised hospital data.^21^ To the best of our knowledge, ICD-PM has never been applied to population-based data collected via VA. This is an important gap due to the significant proportion of home deliveries and deaths, as well as unreliable health records, where the burden of perinatal deaths is greatest. For ICD-PM to be truly globally comparable, relevant, and useful to perinatal death reduction efforts, reporting on experiences of its application to population-based, VA-derived data is essential.

The primary objective of this study was to determine the underlying causes of stillbirths and neonatal deaths in rural Cambodia, using a population-based pregnancy surveillance system combined with VA and the WHO’s ICD-PM classification system. We also aimed to describe the feasibility, strengths, and limitations of ICD-PM when used with VA-derived data, which has not been previously rigorously examined in population-based, low-resource contexts.

## Methods

This was a prospective observational study covering the whole of Preah Vihear province, in North-Eastern Cambodia, over four years (1^st^ September 2018 to 31^st^ August 2022). This study was nested within the Saving Babies’ Lives (SBL) implementation research programme.^22^ SBL is a cluster-randomised trial to evaluate a package of interventions aiming to reduce perinatal mortality in Preah Vihear province. Population-based pregnancy surveillance identified all births and deaths before 28 days of life. Verbal autopsy was conducted with ICD-PM to classify causes of stillbirths and neonatal deaths. A social autopsy component was later added, the results of which are published separately.

### Study setting

Preah Vihear is a large, rural province comprising 1.6% of Cambodia’s population (254,827 in 2019).^23^ Before this study, birth registration among of children under five was 66.5% in the province, below the national average of 73.3%.^9^ No data were available on birth registration by the end of the neonatal period or on death registration.

Primary healthcare in Cambodia is supported by a network of community health workers (CHWs), typically two per village. As trained laypeople, CHWs cover all 289 villages in Preah Vihear province, each serving approximately 441 people. Although they often track pregnant women in their village informally, CHWs have no formal role in reporting births or deaths, and do not usually attend deliveries.

### Surveillance

A neonatal surveillance system harnessing the existing CHW network was set up to cover all deliveries and deaths before the first four weeks of life. CHWs were trained to identify, take verbal consent, and collect data on all pregnancies ≥28 weeks gestation from their respective village, and then follow up the pregnancies until 28 days of life. All deliveries ≥28 weeks gestation (determined from last menstual period, otherwise birth weight ≥1000 grams was used) of women living in and/or delivering in the study area during the study date range were included.

CHWs used a paper-based case record form (CRF) to collect data on estimated delivery date, birth details (date, place, sex, gestation, birth weight, multiple birth), and pregnancy outcome. Outcomes were defined as stillbirth (baby born with no signs of life), neonatal death (death during the first 28 completed days of life) or neonatal survival. Training and familiarisation with the CRF was provided prior to study start. On a monthly basis, the study team met with CHWs and entered the CRF data into a tablet-based electronic form developed using KoboToolbox.^24^ Overdue outcomes were monitored and the relevant CHW prompted accordingly. Missing and unfeasible data checks were done to allow timely clarification with the relevant CHW. CHWs received $2USD per completed CRF to compensate for time and expenses incurred.

### Additional deaths

Due to resource constraints, we focused on finding deaths missed by the surveillance system. In Cambodia, families are legally required to register births and deaths with their local authority (commune council), but we found few births and no stillbirths or neonatal deaths recorded and so did not pursue this data source further. Instead, we reviewed all facility records in the study area annually for stillbirths and neonatal deaths, and cross-checked these with surveillance data to identify additional deaths. Any newly found cases were followed up by the designated CHW. If the family could not be located, the available facility data were used to record the death.

### Verbal autopsy

A VA was performed by the study team within six months of all stillbirths and neonatal deaths, and physician analysis used to assign cause of death using the ICD-PM classification system.

To collect data, we adapted the 2016 WHO VA questionnaire for stillbirths and neonatal deaths to our context and purpose (Appendix 1).^25,26^ A tablet-based, electronic version of the questionnaire was developed in both English and Khmer (the national language) using KoboToolbox.^24^ CHWs helped organise the interviews, which were held in privacy (mostly in the family home) with at least one parent and/or close family member who was present during the time preceding death. Following informed consent, interviews were conducted by the trained Cambodian study team in Khmer, which is the usual spoken language in the study area except for near the Laos border. Completed VAs were checked weekly for completion and logic to allow for timely clarification as needed.

To determine cause of death from VA data collected, we used dual physician analysis. This is the accepted best method for VA analysis, especially for research studies and settings where many deliveries and deaths occur outside of the formal health system.^27,28^ The trained, consistent coding team comprised three paediatricians: a Cambodian-trained and a UK-trained senior registrar who independently assigned cause of death to all VA cases, and a UK-trained consultant with expertise in tropical medicine and obstetrics who independently assigned cause of death only in cases of discrepancy. All three coders were based in Cambodia prior to and throughout the study, and familiar with the local context. A diagnosis of ‘unclassifiable’ was used when agreement between all three physician coders was not achieved.

### ICD-PM

To classify the causes of stillbirths and neonatal deaths we used the WHO ICD-PM classification system.^19^ We adapted the coding system to our context and purpose (Table 1, Appendix 2). The main adaptation was the addition of a fourth major category for the timing of death: ‘U’ for unknown timing of stillbirth in relation to the onset of labour. Subsequent U sub-categories (U1 to U6) followed the existing ICD-PM codes for antepartum stillbirths and those used in previous studies to ensure they were relevant and comparable.^21,29^ We also developed rules for assigning codes that were difficult to differentiate due to a lack of diagnostics and clinical information, such as birth asphyxia (N4) and hypoxic ischaemic encephalopathy (N5) (Appendix 2).

**Table 1:** ICD-PM codes and descriptions adapted and used to classify causes of stillbirths and neonatal deaths in our study.

| Step 1 |  |  | ICD-PM code | Description |
| --- | --- | --- | --- | --- |
|  | Timing of death |  | U | Unknown timing of death in relation to labour onset, died before delivery. |
|  |  |  | A | Dead before/at the onset of labour, died before delivery. |
|  |  |  | I | Alive at onset of labour, died before delivery. |
|  |  |  | N | Alive at delivery, died before 28 days of life. |
| Step 2 | Timing of death |  | ICD-PM code | Description |
|  | U | Stillbirth of unknown timing | U1 | Congenital malformations, deformations, and chromosomal abnormalities |
|  |  |  | U2 | In utero hypoxia |
|  |  |  | U3 | Infection |
|  |  |  | U4 | Other specified in utero disorder |
|  |  |  | U5 | Disorders related to fetal growth |
|  |  |  | U6 | In utero death of unspecified cause |
|  | A | Antepartum stillbirth | A1 | Congenital malformations, deformations, and chromosomal abnormalities |
|  |  |  | A2 | Infection |
|  |  |  | A3 | Antepartum hypoxia |
|  |  |  | A4 | Other specified antepartum disorder |
|  |  |  | A5 | Disorders related to fetal growth |
|  |  |  | A6 | Antepartum death of unspecified cause |
|  | I | Intrapartum stillbirth | I1 | Congenital malformations, deformations, and chromosomal abnormalities |
|  |  |  | I2 | Birth trauma |
|  |  |  | I3 | Acute intrapartum event |
|  |  |  | I4 | Infection |
|  |  |  | I5 | Other specified intrapartum disorder |
|  |  |  | I6 | Disorders related to fetal growth |
|  |  |  | I7 | Intrapartum death of unspecified cause |
|  | N | Neonatal death | N1 | Congenital malformations, deformations, and chromosomal abnormalities |
|  |  |  | N2 | Disorders related to fetal growth |
|  |  |  | N3 | Birth trauma |
|  |  |  | N4 | Complications of intrapartum events |
|  |  |  | N5 | Convulsions and disorders of cerebral status |
|  |  |  | N6 | Infection |
|  |  |  | N7 | Respiratory and cardiovascular disorders |
|  |  |  | N8 | Other neonatal conditions |
|  |  |  | N9 | Low birth weight and prematurity |
|  |  |  | N10 | Miscellaneous |
|  |  |  | N11 | Neonatal death of unspecified cause |
| Step 3 |  |  | ICD-PM code | Description |
|  | Maternal contributing condition |  | M1 | Complications of placenta, cord and membranes |
|  |  |  | M2 | Maternal complications of pregnancy |
|  |  |  | M3 | Other complications of labour and delivery |
|  |  |  | M4 | Maternal medical and surgical conditions |
|  |  |  | M5 | No maternal condition |

For each case we applied the ICD-PM in three steps (Table 1, Appendix 2):

1. Time of death major category: exact timing of death was classified as antepartum stillbirth (A), intrapartum stillbirth (I), unknown timing of stillbirth (U), or neonatal death (N).
2. Primary cause of death sub-category: for each timing category (A, I, U, N) medical cause of fetal or neonatal death was coded as A1 to A5, I1 to I7, U1 to U6, N1 to N11.
3. Contributing maternal condition category: main maternal condition or disease considered to have contributed to death was coded as M1 to M5.

Only one primary cause of death and one maternal condition was permitted: this was considered to be the earliest condition in the chain of events ending in death. When physician coders did not agree on the primary cause of death, but did agree on the time of death code, only the major timing of death code (U, A, I, N) was reported and the primary cause described as ‘unclassifiable’. When the coders did not agree on the maternal contributing condition, this was also reported as ‘unclassifiable’.

The ICD-PM system is designed to be used for deaths in the perinatal period: all stillbirths and early (within the first seven days) neonatal deaths. However, ICD-PM can also be used for late neonatal deaths as these may also occur as a consequence of perinatal events.^19^ Thus, for deaths assigned a neonatal (N) code, we report their ICD-PM codes split by timing of death, defined as before and after the first seven completed days of life (early and late neonatal deaths respectively).

### Data analysis

All data were entered on KoboCollect^24^ and analysis was performed using the R software package.^30^ Additional deaths discovered from facility records and not from the surveillance system were excluded both from mortality rate calculations due to their unknown denominator, and from comparison of characteristics of deliveries that survived with those that died. Due to few missing data (3.6% for the multivariable analysis), complete case analysis was performed. Causes of deaths were described as cause-specific mortality fractions (CSMFs), the accepted metric in the field.^31^ For physician analysis, acceptable inter-rater variability was predefined as equal to or more than 60% and assessed using Cohen’s Kappa statistic. Descriptive analyses included frequencies, percentages, and rates. Relative risks with 95% confidence intervals were calculated to assess associations between perinatal risk factors and death. Potential selection bias was assessed by comparing characteristics of VA enrolled and non-enrolled cases.

### Ethics approval

The sensitive nature of VA required careful ethical consideration. Verbal autopsy was conducted after allowing for a mourning period, and confidentiality of cases and of participant views was maintained at all times. The study was approved by the Cambodian National Ethics Committee for Health Research (NECHR, 283) and the Oxford Tropical Research Ethics Committee (OxTREC, 547–17). Verbal autopsy interviews were done after obtaining informed verbal consent from caregivers of the deceased.

## Results

There were 23,975 deliveries and 457 deaths (198 stillbirths and 259 neonatal deaths) recorded by the surveillance system during the study (Figure 1). The overall stillbirth rate, neonatal mortality rate, and perinatal mortality rate were 8.3 per 1000 deliveries, 10.9 per 1000 live births, and 17.4 per 1000 deliveries respectively.

**Figure 1.**
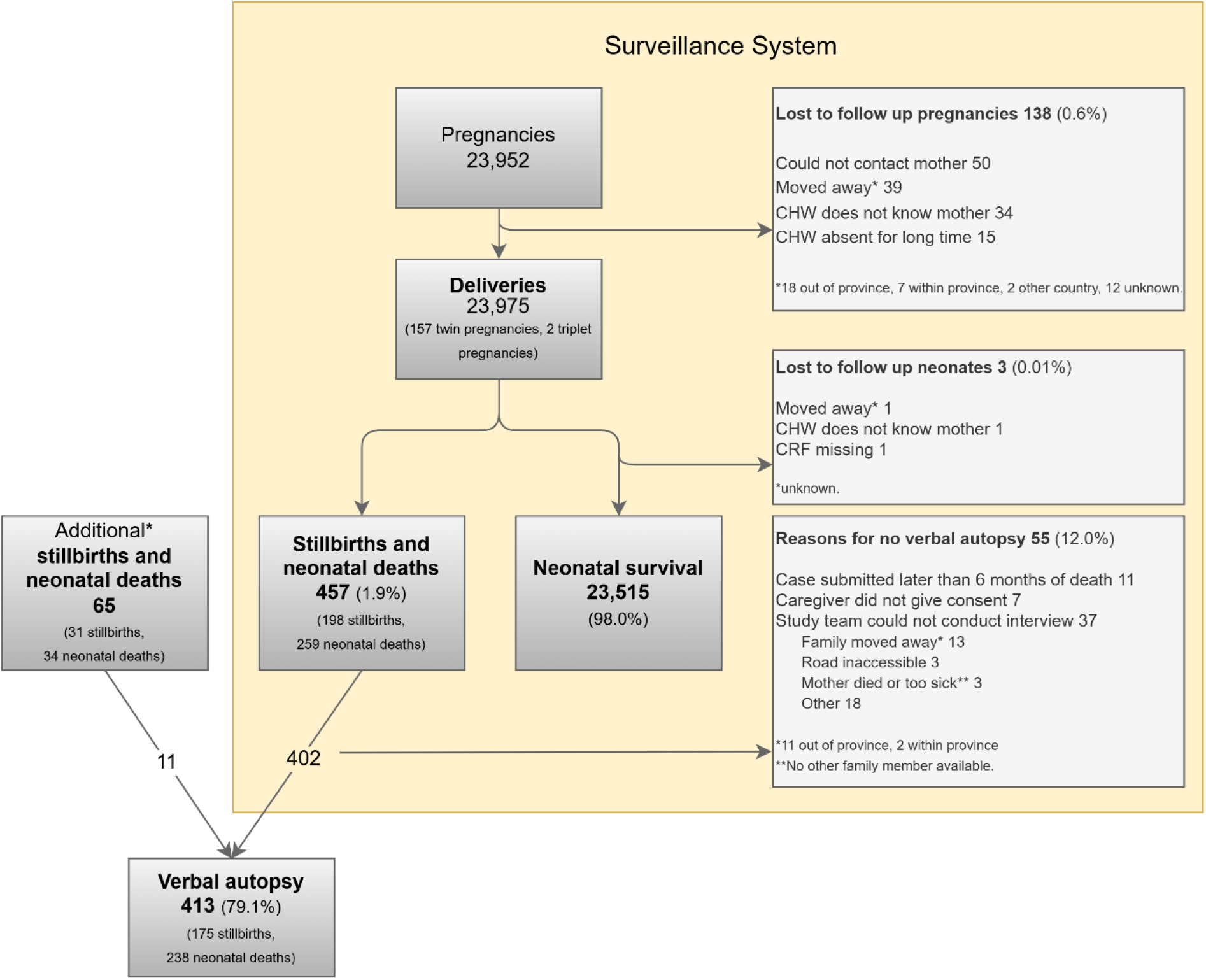
Flowchart of stillbirths and perinatal deaths. Community health worker (CHW); case record form (CRF). *additional death cases found from health facility records.

Baseline characteristics of all deliveries found via surveillance are shown in Table 2. Most deliveries occurred in primary (63.2%) and secondary (21.6%) government-run facilities in the province and 5.5% of deliveries occurred in the community. Of the 522 deaths, 100 (19.2%) occurred in the community. Death (both stillbirths and neonatal deaths) was associated with male sex, preterm (<37 weeks gestation) delivery, low birth weight, and being born in the community (Appendix 3).

**Table 2.** Characteristics of deliveries ≥28 weeks gestation, stratified by outcome: Deceased (stillbirths and neonatal deaths) or neonatal survival. Birth location definitions: Enroute: deliveries that occurred whilst travelling to a facility; Primary: primary-level government healthcare facility; Secondary: government hospital; Private: privately-run health facility; Tertiary: tertiary-level non-governmental organisation (NGO)-run hospital. Birth locations were all in the study area (Preah Vihear province) unless stated as Other province.

| Characteristic | Deceased<br>N = 457 | Neonatal survival<br>N = 23,515 | Overall<br>N = 23,972 |
| --- | --- | --- | --- |
| **Male sex, n (%)** | 278 (61.2%) | 12,175 (51.8%) | 12,453 (52.0%) |
| Missing | 3 | 0 | 3 |
| **Preterm\*, n (%)** | 232 (50.9%) | 717 (3.1%) | 949 (4.0%) |
| Missing | 1 | 34 | 35 |
| **Birth weight (kg), median (IQR)** | 2.30 (1.50, 3.00) | 3.00 (2.80, 3.30) | 3.00 (2.80, 3.30) |
| Missing | 88 | 758 | 846 |
| **Multiple birth, n (%)** | 28 (6.1%) | 283 (1.2%) | 311 (1.3%) |
| Missing | 0 | 0 | 0 |
| **Birth location, n (%)** |  |  |  |
| Home | 54 (11.8%) | 1,266 (5.4%) | 1,320 (5.5%) |
| Enroute | 22 (4.8%) | 0 (0.0%) | 22 (0.1%) |
| Primary | 170 (37.2%) | 14,973 (63.7%) | 15,143 (63.2%) |
| Secondary | 150 (32.8%) | 5,029 (21.4%) | 5,179 (21.6%) |
| Private | 6 (1.3%) | 384 (1.6%) | 390 (1.6%) |
| Other | 2 (0.4%) | 67 (0.3%) | 69 (0.3%) |
| Other province Primary | 6 (1.3%) | 437 (1.9%) | 443 (1.8%) |
| Other province Secondary | 7 (1.5%) | 342 (1.5%) | 349 (1.5%) |
| Other province Tertiary (NGO) | 38 (8.3%) | 1,012 (4.3%) | 1,050 (4.4%) |
| Other province Private | 2 (0.4%) | 0 (0.0%) | 2 (0.0%) |
| Missing | 0 | 5 | 5 |

\*Preterm defined as <37 weeks gestational age

### Deaths

Checking health facility records found 155 (29.7%, 155/522) stillbirths and neonatal deaths, of which 65 had not yet been identified by the surveillance system. Of these 65 additional deaths, data could be collected by CHWs for 30 cases, and 11 had a VA. Some differences were found in baseline characteristics between the 457 cases found by the CHW surveillance system and the additional 65 cases found from facility records (Appendix 4).

Overall 522 deaths (229 stillbirths and 293 neonatal deaths) were found. For stillbirth cases with a known timing of death, 61.0% (94/154) of stillbirths occurred antepartum (Figure 2). The proportion of neonatal deaths occurring on the first day of life was 41.6% (122/293) and in the first week of life was 86.0% (252/293) (Figure 2).

**Figure 2.**
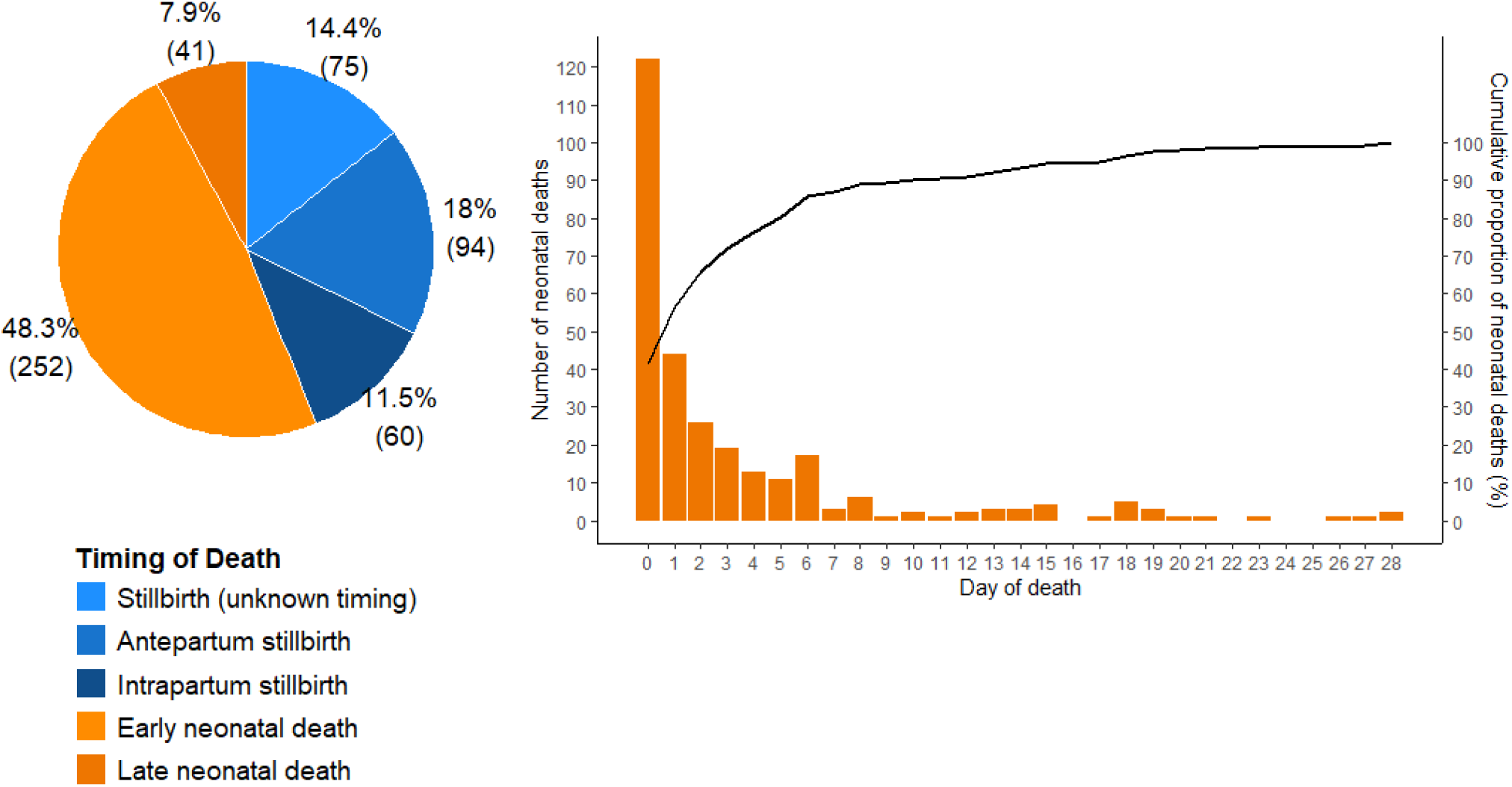
Death cohort (n=522): timing of deaths.

Baseline characteristics of all deaths, stratified by the timing of their death, are shown in appendix 5. Amongst the 522 deaths, 15.3% occurred in the community.

### Verbal autopsy

Verbal autopsy was conducted on 413 of the 522 deaths (79.1%). Reasons for non-enrollment included deaths being reported too late (> six months), lack of consent, and logistical issues including road access issues and the family moving away (Figure 1). We found no difference between the baseline characteristics of the VA enrolled and unenrolled population, except for in birth and death locations (Appendix 6). The median time from the date of death to VA interview was 54.0 days (IQR 30.0-91.0 days). By the time of VA interview, five mothers had died (two intrapartum and three postpartum). Excluding these five maternal death cases, the mother was the main respondent for 98.5% (402/408) of cases.

For 67 of the 413 (16.2%) VA cases no agreement on cause of death could be reached between the initial two physician analysers and so the case went to a third physician. Of these 67 cases, an agreement could be reached between two out of the three physicians for 44 cases. For all 23 cases for which physician agreement was not reached on the primary cause of death (unclassifiable), an agreement was reached on the timing of death and so the major code is reported (table 3). Inter-rater reliability between the two primary physicians conducting physician analysis was 0.811, calculated using Cohen’s Kappa statistic.

**Table 3:** ICD-PM coding of stillbirths and neonatal deaths.

| Maternal contributing condition |  |  |  |  |  |  |  |  |
| --- | --- | --- | --- | --- | --- | --- | --- | --- |
|  |  | M1 | M2 | M3 | M4 | M5 | M | Total |
|  |  | Complications of placenta, cord and membranes | Maternal complications of pregnancy | Other complications of labour and delivery | Maternal medical and surgical conditions | No maternal condition | Unclassifiable |  |
| Unknown timing of stillbirth |  |  |  |  |  |  |  |  |
| U1 | Congenital malformations | 0 (0.0%) | 0 (0.0%) | 0 (0.0%) | 0 (0.0%) | 1 (4.8%) | 0 (0.0%) | 1 (4.8%) |
| U6 | Unspecified cause | 0 (0.0%) | 0 (0.0%) | 0 (0.0%) | 1 (4.8%) | 6 (28.6%) | 0 (0.0%) | 7 (33.3%) |
| U | Unclassifiable | 2 (9.5%) | 0 (0.0%) | 0 (0.0%) | 5 (23.8%) | 6 (28.6%) | 0 (0.0%) | 13 (61.9%) |
| Subtotal |  | 2 (9.5%) | 0 (0.0%) | 0 (0.0%) | 6 (28.6%) | 13 (61.9%) | 0 (0.0%) | 21 (100.0%) |
| Antepartum stillbirth |  |  |  |  |  |  |  |  |
| A1 | Congenital malformations | 0 (0.0%) | 0 (0.0%) | 0 (0.0%) | 0 (0.0%) | 2 (2.1%) | 0 (0.0%) | 2 (2.1%) |
| A2 | Infection | 1 (1.1%) | 0 (0.0%) | 0 (0.0%) | 4 (4.3%) | 0 (0.0%) | 0 (0.0%) | 5 (5.3%) |
| A3 | Antepartum hypoxia | 5 (5.3%) | 0 (0.0%) | 0 (0.0%) | 1 (1.1%) | 1 (1.1%) | 0 (0.0%) | 7 (7.4%) |
| A4 | Other condition | 0 (0.0%) | 0 (0.0%) | 0 (0.0%) | 1 (1.1%) | 0 (0.0%) | 0 (0.0%) | 1 (1.1%) |
| A5 | Disorders related to fetal growth | 0 (0.0%) | 0 (0.0%) | 0 (0.0%) | 1 (1.1%) | 0 (0.0%) | 0 (0.0%) | 1 (1.1%) |
| A6 | Unspecified cause | 2 (2.1%) | 3 (3.2%) | 0 (0.0%) | 5 (5.3%) | 66 (70.2%) | 1 (1.1%) | 77 (81.9%) |
| A | Unclassifiable | 0 (0.0%) | 0 (0.0%) | 0 (0.0%) | 1 (1.1%) | 0 (0.0%) | 0 (0.0%) | 1 (1.1%) |
| Subtotal |  | 8 (8.5%) | 3 (3.2%) | 0 (0.0%) | 13 (13.8%) | 69 (73.4%) | 1 (1.1%) | 94 (100.0%) |
| Intrapartum stillbirth |  |  |  |  |  |  |  |  |
| I3 | Intrapartum hypoxia | 4 (6.7%) | 4 (6.7%) | 37 (61.7%) | 1 (1.7%) | 0 (0.0%) | 1 (1.7%) | 47 (78.3%) |
| I7 | Unspecified cause | 0 (0.0%) | 0 (0.0%) | 1 (1.7%) | 1 (1.7%) | 9 (15.0%) | 0 (0.0%) | 11 (18.3%) |
| I | Unclassifiable | 0 (0.0%) | 0 (0.0%) | 0 (0.0%) | 1 (1.7%) | 1 (1.7%) | 0 (0.0%) | 2 (3.3%) |
| Subtotal |  | 4 (6.7%) | 4 (6.7%) | 38 (63.3%) | 3 (5.0%) | 10 (16.7%) | 1 (1.7%) | 60 (100.0%) |
| Stillbirth subtotal |  | 14 (8.0%) | 7 (4.0%) | 38 (21.7%) | 22 (12.6%) | 92 (52.6%) | 2 (1.1%) | 175 (100.0%) |
| Early neonatal death |  |  |  |  |  |  |  |  |
| N1 | Congenital malformations | 0 (0.0%) | 1 (0.5%) | 0 (0.0%) | 1 (0.5%) | 11 (5.4%) | 1 (0.5%) | 14 (6.9%) |
| N4 | Intrapartum hypoxia | 10 (4.9%) | 3 (1.5%) | 32 (15.8%) | 0 (0.0%) | 13 (6.4%) | 3 (1.5%) | 61 (30.0%) |
| N6 | Infection | 0 (0.0%) | 0 (0.0%) | 3 (1.5%) | 4 (2.0%) | 24 (11.8%) | 3 (1.5%) | 34 (16.7%) |
| N8 | Other condition | 0 (0.0%) | 0 (0.0%) | 0 (0.0%) | 0 (0.0%) | 1 (0.5%) | 0 (0.0%) | 1 (0.5%) |
| N9 | Low birth weight and prematurity | 9 (4.4%) | 4 (2.0%) | 59 (29.1%) | 4 (2.0%) | 3 (1.5%) | 4 (2.0%) | 83 (40.9%) |
| N11 | Unspecified cause | 0 (0.0%) | 0 (0.0%) | 0 (0.0%) | 0 (0.0%) | 3 (1.5%) | 0 (0.0%) | 3 (1.5%) |
| N | Unclassifiable | 0 (0.0%) | 0 (0.0%) | 1 (0.5%) | 1 (0.5%) | 4 (2.0%) | 1 (0.5%) | 7 (3.4%) |
| Subtotal |  | 19 (9.4%) | 8 (3.9%) | 95 (46.8%) | 10 (4.9%) | 59 (29.1%) | 12 (5.9%) | 203 (100.0%) |
| Late neonatal death |  |  |  |  |  |  |  |  |
| N1 | Congenital malformations | 0 (0.0%) | 0 (0.0%) | 0 (0.0%) | 0 (0.0%) | 6 (17.1%) | 0 (0.0%) | 6 (17.1%) |
| N4 | Intrapartum hypoxia | 0 (0.0%) | 0 (0.0%) | 0 (0.0%) | 0 (0.0%) | 1 (2.9%) | 0 (0.0%) | 1 (2.9%) |
| N6 | Infection | 0 (0.0%) | 0 (0.0%) | 2 (5.7%) | 1 (2.9%) | 15 (42.9%) | 0 (0.0%) | 18 (51.4%) |
| N9 | Low birth weight and prematurity | 0 (0.0%) | 3 (8.6%) | 4 (11.4%) | 0 (0.0%) | 0 (0.0%) | 1 (2.9%) | 8 (22.9%) |
| N11 | Unspecified cause | 0 (0.0%) | 0 (0.0%) | 0 (0.0%) | 0 (0.0%) | 1 (2.9%) | 0 (0.0%) | 1 (2.9%) |
| N | Unclassifiable | 0 (0.0%) | 0 (0.0%) | 0 (0.0%) | 0 (0.0%) | 1 (2.9%) | 0 (0.0%) | 1 (2.9%) |
| Subtotal |  | 0 (0.0%) | 3 (8.6%) | 6 (17.1%) | 1 (2.9%) | 24 (68.6%) | 1 (2.9%) | 35 (100.0%) |
| Neonatal subtotal |  | 19 (8.0%) | 11 (4.6%) | 101 (42.4%) | 11 (4.6%) | 83 (34.9%) | 13 (5.5%) | 238 (100.0%) |
| Total |  | 33 (8.0%) | 18 (4.4%) | 139 (33.7%) | 33 (8.0%) | 175 (42.4%) | 15 (3.6%) | 413 (100.0%) |

### Causes of deaths

Final CSMFs classified by ICD-PM codes are shown in Table 3 and Figure 3. For 12.0% (21/175) of stillbirths the timing of the death in relation to the onset of labour could not be determined (U). No cause of death could be identified for 95.2% (20/21) of stillbirths of unknown timing, 83.0% (78/94) of antepartum stillbirths, but only 21.7% (13/60) of intrapartum stillbirths. For intrapartum stillbirths, hypoxia (78.3%, 47/60) was the leading (and only) cause identified.

**Figure 3.**
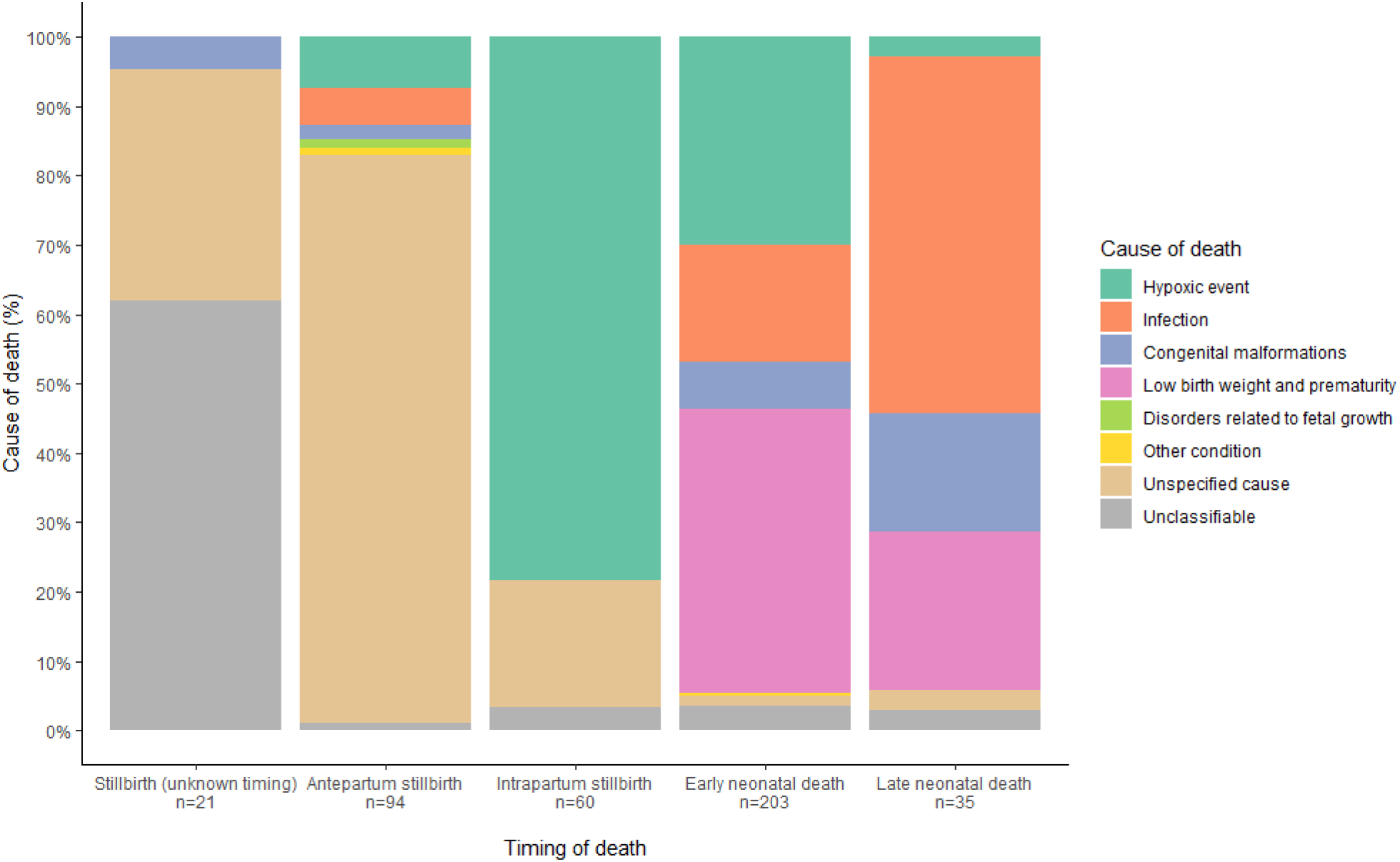
Cause-specific mortality fractions by timing of death.

Most deaths occurred intrapartum and within the first seven days of life (63.7% (263/413)). During this critical time period the leading cause of death and associated maternal condition were hypoxia (41.1%, 108/263), and complications of labour and delivery (50.6%, 133/263) respectively (Appendix 7).

Four causes (low birth weight and prematurity, intrapartum hypoxia, infection, and congenital malformations) accounted for nearly all (94.5%, 225/238) neonatal deaths. Most early neonatal deaths were attributed to low birth weight and prematurity (40.9%, 83/203) and intrapartum hypoxia (30.0%, 61/203), and complications of labour and delivery (46.8%, 95/203) was the commonest maternal contributing condition. Most late neonatal deaths were attributed to infection (51.4%, 18/35), and only one case was attributed to hypoxia (2.9%); maternal contributing cause was largely unattributed (68.6%, 24/35). The CSMFs for neonatal deaths changed with progressing age, such that after 19 days of life, the only causes were infection and congenital abnormalities (Appendix 8).

## Discussion

Our study provides the most comprehensive description of perinatal epidemiology in Cambodia. To the best of our knowledge, it is also the first reported study combining VA with ICD-PM for both stillbirths and neonatal deaths in both community and facility (all) settings.

Our stillbirth and neonatal mortality rates are lower than recently reported in Cambodia,^14^ although the perinatal mortality is the same as that reported in Preah Vihear province in the most recent Cambodian DHS.^23^

In keeping with global data, preterm and low birth weight complications were the leading cause of neonatal deaths in our setting.^32^ The only previous neonatal VA study in Cambodia found sepsis to be the leading cause, followed by asphyxia, but this study was based on only 12 neonatal deaths.^13^ Our findings confirm the emphasis on preterm labour as a global target to reduce perinatal mortality.^33,34^

Primary cause of death could not be ascertained for the majority of antepartum stillbirths and stillbirths of unknown timing, similar to experiences applying ICD-PM in other LMICs.^29^ Identifying causes of antepartum deaths in low-resource settings remains a global challenge as without this information, interventions cannot be targeted.^35^

### Strengths & Limitations: Surveillance system

Community health worker surveillance of pregnancies was far superior to routinely-collected facility data at discovering deaths. Of all deaths, 84.7% occurred in a facility. However, had we relied solely on facility records, we would only have found 29.7% of deaths, suggesting that facilities were not recording these deaths. This is an important consideration for programmes in similar settings that use routinely-collected mortality data. However, the CHW surveillance system was not perfect: if we had only used surveillance, we would only have found 87.5% (457/522) of all deaths because it missed 65 deaths. Of the 522 deaths, 35 (6.7%) were never found by CHWs and only facility record data were used. Reasons for CHWs missing death cases include the movement of domestic economic migrants in and out of the province, families living in very remote farms for work, and concealment of pregnancy by unmarried women. Future programmes should consider triangulation of data if resources allow, and ideally should also include facility births to provide calculation of more comprehensive mortality rates.

A strength of our surveillance system was harnessing the existing CHW network. Community health workers live amongst the communities they serve, making them accessible and trusted members of the health system.^36^ This might help to explain our high follow-up rate, high proportion of VA enrollment, and high reported home birth rate of 5.5% compared to 3.6% reported in Preah Vihear province.^10^ Our study highlights the importance of CHWs as an important resource in LMICs, particularly in rural and remote settings, to achieve inclusion of the most marginalised communities.

However, the challenge was the limited capacity for data collection as CHWs are busy people with work, family life, and other CHW responsibilities. For the surveillance system, therefore, we prioritised data quality and completeness of a few, core variables, which limited comparison between deceased and surviving babies. Also, lack of sociodemographic data limited our ability to assess the generalisability of results from the enrolled to the unenrolled population.

Another limitation was the lack of early, accurate, dating ultrasound scan results, which meant gestational age (≥28 weeks gestation) for study inclusion could not be verified.

The other major challenge of the pregnancy surveillance system was it’s resource-intensity, making it difficult to be sustained without significant funding, technical, and logistical support. Considerable resources are required to report truthful, accurate data in low-resource settings until routine data collection systems improve.

Finally, many VA unenrolled cases involved additional deaths found from facility records and initially missed by CHWs. Actively including health facility staff in our surveillance system might have resulted in reporting of these deaths sooner and improved our VA enrollment rate.

### Strengths & Limitations: Verbal autopsy

Verbal autopsy is a resource-intensive epidemiological tool, especially combined with dual physician analysis. Another limitation of VA is its reliance solely on observation, perspective, and recall of family members present at the time of terminal illness. Potential for bias towards visible, untreated, and memorable pathology (such as visible congenital abnormalities and prematurity), especially when diagnostics and reliable medical records are lacking, might explain our limited variety of causes of death: we used only 18 out of 30 possible ICD-PM codes.

Antepartum stillbirths are particularly challenging, since diagnostics are needed for usually covert pathology, which might explain the high proportion (70.2%) of antepartum stillbirths with neither primary cause of death, nor maternal condition identified in our setting. Also, without diagnostics (antenatally and/or postnatally), the clinical manifestation of ‘invisible’ pathology might have led to misclassification. For example, a congenital cardiac defect in a term neonate manifesting as respiratory distress might have been misdiagnosed as infection. However, our findings are consistent with those from other LMICs.^29^

Additionally, 98.5% of respondents were mothers and with most deaths occuring around the time of delivery, their own condition might have affected their observation of events. Furthermore, perspectives can be context-specific; for example, neonatal jaundice is a poorly recognised medical problem in rural Cambodia.^37^

Lack of a ‘true’ gold standard to assess accuracy and validate our VA results is another limitation. Triangulation of data with healthcare worker perspectives was not feasible. However, VA as a tool has been validated, although there is less reported experience with stillbirths.^18^ Our findings are comparable to other studies in rural areas, including a multi-country VA study for stillbirths and early neonatal deaths.^38^

### Strengths & limitations: ICD-PM

The ICD-PM system comprises the capacity to account for the unique set of circumstances that occur perinatally: firstly, the transition from the in utero fetal period to the ex utero neonatal period, and secondly the inextricable link between mother and baby. It summarises the complexity of perinatal causes of deaths into a concise four-character code (eg. N9-M3).

We found the multilayered nature of the ICD-PM system to be a great strength because it allowed for classification from a broad to a more detailed level, according to the data available. This meant we could assign ICD-PM to all deaths, even when information from VA was limited or when physicians agreed on the major timing of death code (U/A/I/N) but disagreed on the primary cause of death code (unclassifiable). This maximised the number of cases contributing to perinatal death data, even if this was only timing of death.

We found linking deaths to maternal conditions to be a strength of ICD-PM, particularly in cases when the primary cause of death was unknown but a maternal contributing condition was identified. Thus, the code could still provide useful data to guide public health interventions.

For this study, given the limited availability of detailed information via VA, we accept the results of fewer and broader cause of death categories with relatively high proportions of unknowns. We do not consider a high proportion of unknowns for each of the three steps of the ICD-PM, and especially for antepartum stillbirths, to be a failure of the classification system, but rather a reflection of the context.

### ICD-PM adaptation for use with verbal autopsy

The ICD-PM was developed to be applicable globally but wasn’t piloted in a low-income setting, despite most deaths occuring here.^39^ Subsequent application globally has been mostly on facility data, mostly retrospective, and predominantly in high- and middle-income countries.^21^ We agree with the limited studies from low-resource settings that to be applicable globally, revisions to ICD-PM are both necessary and useful.^21,29^ Based on our experience of using ICD-PM with VA data to identify cause of death, our suggested amendments to the classification system are summarised below.

Most importantly, ICD-PM needs to have the capacity to account for an indeterminate timing of death. The step-wise application of ICD-PM means that without a major timing code, the subsequent cause of death code, even if known, cannot be assigned. As a result these cases would otherwise be excluded from global perinatal death reporting. In our study, 12.0% (21/175) of stillbirth cases were assigned a U code for unknown timing of stillbirth. Even when using hospital records, which might, for example, have documentation on intrapartum heart rate, a multi-country LMIC study assigned a U code to 7.3% (92 cases) of stillbirths.^29^ Since with VA methodology we relied solely on recall from family members, a higher U proportion is to be expected, and emphasises the need for addition of this code to the ICD-PM to make it applicable to VA-derived data. Of note, it can also be very challenging to differentiate between an intrapartum stillbirth and neonatal death within a few minutes of life, especially based on VA data and/or for home deaths.^40^ The ICD-PM also does not account for this unknown time period. Although we did not encounter the need for such a code in our study, it is worth considering for future ICD-PM revisions.

Finally, simplification of ICD-PM codes is a worthwhile consideration for future application to VA-derived data. We developed internal coding rules (Appendix 2) to reduce ambiguity and ensure consistency.

### Implications for practice, policy, and research

Our findings indicate key intervention areas to reduce preventable perinatal deaths. These include promoting facility-based deliveries and strengthening healthcare delivery, particularly obstetric care at delivery, neonatal resuscitation, and care of small and preterm neonates.

The resource-intensity of our pregnancy surveillance system highlights the challenges of generating high-quality, community-level data to capture all births and deaths in low-resource settings. Leveraging existing community-based networks, especially those trusted by the community, can be invaluable in capturing information that would otherwise go unrecorded.

Adaptation of ICD-PM to our context was crucial for meaningful and consistent causes of death assignment. Future application of ICD-PM to population-based, VA data should similarly consider context-specific adaptations.

## Conclusion

The ICD-PM classification system can be applied to VA-derived data but only with adaptation. In particular, the capacity to account for unknown timing of stillbirth was crucial. With adaptation to our context, we found ICD-PM user-friendly, optimised inclusion of cases, and provided a comprehensive description of causes of deaths. Thus, our findings provide globally-comparable, relevant and practical data for programme design and policy makers. To improve the standardisation of perinatal cause of death reporting in regions with the highest mortality rates, important ICD-PM revisions to make it better applicable in LMICs and with VA-derived data are needed.

## Data Availability

The datasets generated and/or analysed during the current study are not publicly available due to ethical and legal reasons, including participant anonymity and privacy, but may be available from the corresponding author on reasonable request. A data access agreement will be put in place prior to data transfer.

## Abbreviations

LMIC: low- and middle-income country
CRVS: civil registration and vital statistics
DHS: demographic health survey
VA: verbal autopsy
ICD-PM: International Classification of Diseases to Perinatal Mortality
SBL: Saving Babies’ Lives
CHW: community health worker
CRF: case record form
CSMF: cause-specific mortality fraction
NECHR: National Ethics Committee for Health Research
OxTREC: Oxford Tropical Research Ethics Committee

## Contributors

KP and CT conceptualised and designed the study. KP and CT developed the methodology. KP and AR developed the software. KP, DL, SK, SD, CL and KL conducted data collection. KP, AR and DL conducted data curation. KP, SP and CT conducted physician analysis. KP conducted data analysis and data visualisation. KP drafted the initial manuscript. KP, CT and VC reviewed the results, interpreted findings, and edited the manuscript. SP translated the abstract. CT and the Angkor Hospital for Children fundraising team acquired funding. KP and DL managed and coordinated the project. CT and VC provided supervision. All authors reviewed and approved the final version of the manuscript.

## Data Sharing Statement

The datasets generated and/or analysed during the current study are not publicly available due to ethical and legal reasons, including participant anonymity and privacy, but may be available from the corresponding author on reasonable request. A data access agreement will be put in place prior to data transfer. Instructions and the data application form are available here: https://www.tropmedres.ac/units/moru-bangkok/bioethics-engagement/data-sharing.

## Acknowledgements

The authors wish to thank all community health workers, the Preah Vihear Provincial Health Department, and the Cambodian Ministry of Health for their support of this study. We also thank staff from Angkor Hospital for Children involved with this study, particularly Dr Ngoun Chanpheaktra, Mara Kik, Lorn Loeuk, Dary Vanna, Khemrouth Cheam, Gabriella Watson, and the fundraising team including Patrick Davis. Finally, we thank all women, babies, and their families for their participation, particularly those that were bereaved.

